# Executive Functioning and Processing Speed as Predictors of Global Cognitive Decline in Alzheimer Disease

**DOI:** 10.1101/2024.10.31.24316508

**Authors:** John P. Haran, A M Barrett, YuShuan Lai, Samuel N. Odjidja, Protiva Dutta, Patrick M McGrath, Imane Samari, Lethycia Romeiro, Abigail Lopes, Vanni Bucci, Beth A. McCormick

## Abstract

**INTRODUCTION:** There is a lack of cognitive tools to predict disease progression in mild cognitive impairment (MCI) and Alzheimer’s disease (AD).

**METHODS:** We assessed patients with MCI, AD, and cognitively healthy controls (cHC) using NIH toolbox assessments for attention/concentration and executive functioning and overall cognitive decline by the Alzheimer’s Disease Assessment Scale-Cognitive (ADAS-Cog).

**RESULTS:** Among 183 participants over a median follow-up of 540 days, both between- and within-subjects variance in NIH toolbox and ADAS-Cog assessments increased from cHC to MCI to AD patients. Among patients with AD, pattern comparison processing speed (PCPS) and dimensional change card sort tests (DCCS) declined at 3 and 6 months prior to global cognitive decline (p=0.008 & 0.0012). A 5-point decrease in either PCPS or DCCS increased risk of global cognitive decline (HR 1.32 (1.08-1.60) and 1.62 (1.16-2.26)).

**DISCUSSION:** Testing for cognitive domains of attention/concentration and executive functioning may predict subsequent global cognitive, and functional decline.

## INTRODUCTION

Today there are nearly 6.9 million older Americans living with Alzheimer’s disease (AD) (1). AD is a progressive neurodegenerative disorder, however, there is much variability in the observed rates of cognitive decline across the spectrum of AD (2–4). The average duration of the disease varies between 4 and 8 years, with some upwards of 20 years (5). There are known demographic and clinical characteristics as well as radiological and genetic features that associate with the rate of cognitive decline (6, 7). More recently both central and peripheral biomarkers have been identified as predicting long-term cognitive decline (8–10). However, these risk factors do not provide short-term predictions of AD disease trajectory. The ability to predict the timing of when a patient will become completely dependent on others would be a powerful tool, enabling patients and families to optimize care and, with newer disease-modifying therapeutics, possibly optimize timing of therapeutic interventions.

The sequence of deficits affecting different cognitive domains in AD commonly starts with an individual’s executive functioning and processing speed (11). Executive functioning defines the higher-level cognitive skills used to control other cognitive abilities (12, 13) while processing speed is the time it takes to execute multi-step information processing in a mental task (14). Both executive functioning and processing speed support other cognitive processes (15, 16) and may serve as a useful cognitive markers for the early trajectory of AD symptoms, being potentially predictive of a more rapid decline (17, 18). Thus, both of these domains may serve as predictors of an inflection point in AD symptoms, when the individual might begin a period of more rapid global cognitive decline, losing function and freedom.

In current clinical practice, there are no established guidelines or tools for monitoring cognitive function after diagnosis for the sole purpose of predicting decline. We reasoned that given that deficits in executive function and processing speed often precede more severe cognitive deficits, standardized testing of these two domains may hold value in clinical practice. While not specifically developed for older populations, the U.S. National Institutes of Health Toolbox for the Assessment of Neurological and Behavioral Function (NIH toolbox) includes a Cognition Battery (CB) that includes brief, comprehensive tests for these functions (19). This CB contains a Dimensional Change Card Sort (DCCS) test that is a sensitive and reliable measure of executive functioning and cognitive flexibility (20), and a Pattern Comparison Processing Speed Test (PCPS), which can also evaluate attention and concentration (21). These NIH toolbox assessments are easy to administer and offer standardized, non-invasive measures which can be compared across studies (19).

The aim of this interim analysis of the Gut-brain Alzheimer’s disease Inflammation and Neurocognitive Study (GAINS) cohort was to evaluate early cognitive data as potential predictors of AD-associated global cognitive decline as measured by the modified Alzheimer’s Disease Assessment Scale-Cognitive Subscale 13 (ADAS-Cog13). ADAS-Cog is a well-established standard for the assessment of cognitive function in patients with AD with moderate to severe disease and is routinely used to help diagnose patients as having MCI, AD, or normal cognition (22) as well as to measure clinically relevant changes (23, 24). The addition of executive functioning and functional ability items to create ADAS-Cog13 (25) from the original 11 question set (ADAS-Cog11) has improved the test’s sensitivity in milder disease (26), but the test still lacks an assessment of processing speed and skews towards language and verbal memory tasks. While greater changes in ADAS-Cog and ADAS-Cog13 scores over time have been associated with AD versus MCI (27), both tests remain in essence, a measure of cognitive ability at a given point in time.

We followed participants in the GAINS cohort at 3-month intervals and assessed their NIH toolbox DCCS and PCPS and ADAS-Cog13 scores. In this longitudinal cohort, a 5-point decrease in the cognitive domains of DCCS or PCPS was predictive of global cognitive and functional decline as measured by the ADAS-Cog13.

## METHODS

### Study Setting and Population

Older adults, ≥60 years of age, living independently, were recruited into the Gut-brain Alzheimer’s disease Inflammation and Neurocognitive Study (GAINS) and included those diagnosed with Alzheimer’s disease (AD), amnestic mild cognitive impairment (MCI), or had no cognitive issues, serving as healthy controls (cHC). Subjects were included in this sub-group analysis if they had completed 4 study visits (270 days). This prospective cohort study was approved by the institutional review board at the University of Massachusetts Chan Medical School.

### Data Collection

We collected demographic measures at enrollment, which included age, sex, race, ethnicity, level of education, and past medical history. At enrollment and at each subsequent visit, we collected information on nutritional status, frailty, and any hospitalizations or changes in medication. We assessed nutritional status using the Mini Nutritional Assessment (MNA) tool (28–30). Subjects were categorized as normal, at risk, or malnourished based on the MNA. Frailty was categorized according to the validated and widely-utilized Canadian Study of Health and Aging’s (CSHA) 7-point Clinical Frailty Scale (31).

During each visit, cognitive testing was performed. We used the modified Alzheimer’s Disease Assessment Scale-Cognitive Subscale 13 (ADAS-Cog13). ADAS-Cog was developed to measure cognitive dysfunction in AD, but is now used for assessment of cognition in other dementia or pre-dementia populations, and is one of the most widely-used cognitive scales in clinical trials (27, 32). We also utilized the NIH toolbox to assess changes in cognition under the subdomains of attention/concentration, using the Pattern Comparison Processing Speed Test (PCPS), and executive functioning using the Dimensional Change Card Sort (DCCS) (33). The Toolbox PCPS provides a reliable measurement of complex processing speed over the lifespan that is sensitive to neurological insults (34).

For our measure of global cognition, we chose to use the ADAS-Cog13 due to its improved responsiveness as compared with the original ADAS-Cog among people with MCI and early disease course AD, as well as its ability to distinguish different stages of AD (22, 26).

Longitudinally, we used a 4-point change in the ADAS-Cog13 score from the day 0 visit as clinically meaningful, either as an improvement (≤ -4 points) or as a decline ≥ +4 points) (35–37). Those that remained within 4 points of their initial visit ADAS-Cog13 scores were categorized as stable, while those with improvement were categorized as improved, and declining as decline. We chose the first timepoint where a subject had a change in ADAS-Cog13 score of ≤ -4 points as the point of decline and the previous visit as the 3 months prior, and the visit before that as 6 months prior to decline. All other previous timepoints were labeled as baseline for analysis.

Determination of AD diagnosis was made by previous cognitive testing coupled with various neuroimaging techniques performed by the subjects’ own physician. Polypharmacy was defined using the most commonly reported definition of five or more daily medications (38). We subdivided our cohort into 4 age categories for analysis: 1) 65 to 74; 2) 75 to 84; 3) 85 to 94; and 4) ≥95 years of age. This has been previously validated in demonstrating signatures of frailty in the gut microbiota (39–41). All survey data was collected by trained research staff or study physicians. Study data were collected and managed using REDCap electronic data capture tools hosted at the University of Massachusetts Chan Medical School.(42)

### Data Analysis

We used chi-square tests to compare categorical variables, and the student’s independent-samples t-test for continuous variables between patient groups with different outcomes from their baseline data. To test the equality of standard deviations (variance) we used the sdtest package in Stata. Cox proportional hazards models were used to evaluate the NIH neuropsychological measures that predicted time to significant global cognitive decline, as defined by a greater than 4-point increase in ADAS-Cog13 testing, as described above. The hazard ratio indicates the change in risk per 5-unit change in the predictor. For instance, if the hazard ratio for PCPS is 1.57, each 5-point change in PCPS equates to an increases risk by 57%. Two sets of adjusted Cox models were completed each for PCPS and DCCS, where these neuropsychological measures were adjusted for age (in years), sex, education level, frailty, malnutrition, and polypharmacy. The software used for the analyses was Stata, Release 13.1 (StataCorp LLC, College Station, TX) and Prism Release 10 (GraphPad by Dotmatics, Ltd., United Kingdom).

## RESULTS

### Clinical and cognitive scores differed among GAINS cohort patient groups

We enrolled 243 older adults, and at the time of this analysis, 183 (75.3%) completed 4 study visits for a longitudinal length of 270 days and were eligible for the analysis. Of these 183 adults in the GAINS cohort, 131 (71.2%) were cHC while 24 (13.0%) had AD and 29 (15.8%) had MCI. This cohort was predominately white (95.1%) and non-Hispanic (91.8%). Unsurprisingly, adults with AD and MCI were older, with higher frailty and malnutrition scores as well as taking more daily medications as defined by polypharmacy (**Table 1**). This is consistent with what is known about frailty and malnutrition in relation to AD and mild cognitive impairment (43–46). Both frail and pre-frail elders usually have poorer cognitive status (47), and frailty has been linked to the extent of AD pathophysiology (44). Malnutrition is also closely linked to decreased cognitive functioning (43). Not surprisingly, mean score on the ADAS-Cog13 increased from cHC, to MCI and AD subjects, and NIH Toolbox module mean scores decreased from cHC, to MCI and AD subjects (**Table 1**). These results further validate the ADAS-Cog13 in responsiveness between cognitively healthy individuals and those with MCI versus AD. They also demonstrate the utility of the NIH toolbox cognitive modules in distinguishing these populations.

**Table 1:** Clinical characteristics of the GAINS cohort.

| **Table 1: Clinical characteristics of the GAINS cohort** |  |  |  |  |
| --- | --- | --- | --- | --- |
|  | **HC** | **MCI** | **AD** | **p-value** |
| Age * | 70.2 (7.6) | 75.4 (7.2) | 74.3 (5.6) | <0.001 |
| Male | 39 (29.8) | 15 (51.7) | 11 (45.8) | 0.042 |
| Education level | 6.0 (1.5) | 5.8 (1.2) | 5.7 (1.7) | 0.43 |
| Malnutrition Score* | 1.2 (0.5) | 1.3 (0.5) | 1.8 (0.7) | <0.001 |
| CFS * | 2.1 (1.0) | 2.6 (1.0) | 3.8 (1.4) | <0.001 |
| Polypharmacy | 34 (26.0) | 10 (34.5) | 16 (66.7) | <0.001 |
| BMI | 27.7 (6.0) | 26.7 (4.9) | 26.1 (6.3) | 0.44 |
| ADAS-Cog13 | 8.9 (4.6) | 19.4 (9.2) | 36.8 (22.5) | <0.001 |
| PCPS | 93.5 (14.2) | 83.6 (16.4) | 66.2 (20.1) | <0.001 |
| DCCS | 101.2 (10.0) | 96.8 (12.5) | 84.0 (18.3) | <0.001 |
| Data presented as n (%) unless marked with * then presented means (sd). CFS – Clinical Frailty Score; BMI – body mass index; ADAS-Cog13 - Alzheimer's Disease Assessment Scale-Cognitive Subscale 13; PCPS – NIH Toolbox Pattern Comparison Processing Speed Test; DCCS - NIH Toolbox Dimensional Change Card Sort |  |  |  |  |

### Cognitive Outcomes in AD was not correlated with initial cognitive scores

Among the GAINS cohort we noted 3 cognitive trajectory patterns on longitudinal ADAS-Cog13 testing. Using a change in score of +/-4 in ADAS-Cog testing for 2 or more timepoints to categorize outcomes, 48 patients improved their scores over time (26.1%), 119 patients remained stable (64.7), and 17 patients experienced cognitive decline (9.2%). Of those with AD, 14 had cognitive decline (58.3%) while 8 remained stable (33.3%) and 2 improved (8.3%). Among AD patients, there were no significant differences among the 3 cognitive outcomes observed (**Table 2**). Importantly, there were no differences in the day 0 cognitive scores using the ADAS-Cog or NIH toolbox among the AD subjects by cognitive trajectory outcome. In the cHC and MCI patient groups, there were a combined n=5 patients with decline (3.1%), who were older and had worse initial ADAS-Cog testing scores (**Supplemental Table 1**).

**Table 2:** Baseline Characteristics of Alzheimer’s disease patients by cognitive Outcomes.

|  |  |  |  |  |
| --- | --- | --- | --- | --- |
|  | **Improve (2)** | **Stable (8)** | **Decline (14)** | **p-value** |
| Age (years/SD)* | 76.0 (4.2) | 76.5 (4.3) | 72.7 (6.2) | 0.30 |
| Male | 0 (0.1) | 4 (50.0) | 7 (50.0) | 0.40 |
| Education level* | 5.0 (2.8) | 6.1 (1.7) | 5.6 (1.7) | 0.52 |
| Malnutrition Score* | 1.5 (0.7) | 2.0 (0.9) | 1.8 (0.6) | 0.64 |
| CFS* | 3.0 (1.4) | 3.8 (1.8) | 3.9 (1.3) | 0.71 |
| Polypharmacy | 1 (50.0) | 6 (75.0) | 9 (64.3) | 0.77 |
| BMI* | 29.8 (4.9) | 26.0 (5.8) | 25.7 (7.0) | 0.70 |
| ADAS-Cog13* | 31.3 (25.0) | 38.8 (29.9) | 38.3 (19.2) | 0.92 |
| PCPS * | 59.0 (14.1) | 74.3 (19.3) | 60.7 (19.4) | 0.35 |
| DCCS* | 79.0 (19.8) | 94.2 (16.1) | 80.5 (19.3) | 0.33 |
| Data presented as n (%) unless marked with * then presented means (sd). CFS – Clinical Frailty Score; BMI – body mass index; ADAS-Cog13 - Alzheimer's Disease Assessment Scale-Cognitive Subscale 13; PCPS – NIH Toolbox Pattern Comparison Processing Speed Test; DCCS - NIH Toolbox Dimensional Change Card Sort |  |  |  |  |

**Supplemental Table 1:** Characteristics of cHC and MCI patients by cognitive outcomes.

|  |  |  |  |  |
| --- | --- | --- | --- | --- |
|  | **Improve (45)** | **Stable (110)** | **Decline (5)** | **p-value** |
| Age (years/SD)* | 73.1 (6.5) | 70.1 (8.0) | 75.4 (9.8) | 0.042 |
| Male | 19 (42.2) | 33 (46.0) | 2 (40.0) | 0.33 |
| Education level* | 5.9 (1.6) | 6.0 (1.4) | 5.6 (2.1) | 0.76 |
| Malnutrition Score* | 1.2 (0.5) | 1.2 (0.5) | 1.3 (0.5) | 0.98 |
| CFS* | 2.3 (0.9) | 2.1 (1.1) | 2.0 (0.8) | 0.73 |
| Polypharmacy | 13 (28.9) | 29 (26.4) | 2 (40.0) | 0.78 |
| BMI* | 28.0 (6.3) | 27.3 (5.7) | 27.2 (3.6) | 0.82 |
| ADAS-Cog13* | 14.0 (5.2) | 9.2 (5.9) | 22.1 (18.0) | <0.001 |
| PCPS * | 87.7 (13.0) | 93.4 (15.6) | 86.3 (18.4) | 0.08 |
| DCCS* | 101.1 (6.9) | 100.2 (11.8) | 96.0 (6.6) | 0.64 |
| Data presented as n (%) unless marked with * then presented means (sd). CFS – Clinical Frailty Score; BMI – body mass index; ADAS-Cog13 - Alzheimer's Disease Assessment Scale-Cognitive Subscale 13; PCPS – NIH Toolbox Pattern Comparison Processing Speed Test; DCCS - NIH Toolbox Dimensional Change Card Sort |  |  |  |  |

### Significant variance in cognitive testing exists between cognitively healthy older adults and those with MCI and AD

To further characterize the baseline characteristics of our study groups (cHC, MCI, AD), we examined the variance between individuals (between) as well as within-person variance (within) (**Table 3**) as standard deviations are not easy to interpret without a frame of reference (48). Both the between- and within-variance increased from cHC to MCI to AD patients for ADAS-Cog13 testing (**Table 3**). For both the NIH toolbox assessment of PCPS and DCCS, we did not observe differences in within-group variance across groups. This is reflected in the increasing intraclass correlation coefficient as one goes from cHC to MCI to AD among all cognitive tests (**Supplemental Table 2**, all p<0.01). The values in Table 3 indicate that there is considerable within-person variability in each of the measures of cognitive functioning. Moreover, the within-person to between-person variability ratios were somewhat larger for the cHC patients compared to those with MCI or AD mostly due to the near doubling of between variance as we move from HC to MCI to AD across all cognitive testing types. The variance for the cHC and MCI groups indicate that the variation for a given individual from one test occasion to the next is more than half as much as the variation from one person to the next. This was the opposite tendency for AD patients where variation for the individual was one quarter to one third of the variation from one person to the next across cognitive tests.

**Table 3:** Variance among different cognitive group types.

| **Table 3: Variance among different cognitive group types** |  |  |  |
| --- | --- | --- | --- |
|  | **HC** | **MCI** | **AD** |
| **ADAS-Cog13** |  |  |  |
| Mean (SD) | 7.5 (4.5) | 17.0 (10.4) | 37.8 (23.3) |
| Between variance | 3.9 | 10.7 | 23.4 |
| Within variance | 2.5 | 3.1 | 5.7 |
| Within/Between | 0.64 | 0.29 | 0.24 |
| **PCPS** |  |  |  |
| Mean (SD) | 99.6 (14.8) | 89.9 (17.8) | 70.6 (20.5) |
| Between variance | 12.5 | 16.7 | 21.4 |
| Within variance | 8.1 | 8.0 | 8.1 |
| Within/Between | 0.65 | 0.50 | 0.38 |
| **DCCS** |  |  |  |
| Mean (SD) | 103.0 (8.8) | 99.7 (10.8) | 87.7 (17.1) |
| Between variance | 7.2 | 10.1 | 19.4 |
| Within variance | 5.3 | 5.5 | 6.1 |
| Within/Between | 0.74 | 0.54 | 0.31 |
| **P-value** | < 0.01 | < 0.01 | < 0.01 |
| HC – Healthy Control; MCI – Mild Cognitive Impairment; AD – Alzheimer’s disease; ADAS-Cog13 - Alzheimer’s Disease Assessment Scale-Cognitive Subscale 13; PCPS – NIH Toolbox Pattern Comparison Processing Speed Test; DCCS - NIH Toolbox Dimensional Change Card Sort |  |  |  |

**Supplemental Table 2:** Intraclass correlation coefficients among different cognitive group types.

|  | HC |  | MCI |  | AD |  |
| --- | --- | --- | --- | --- | --- | --- |
| ADAS-Cog13 | 0.63 | (0.56-0.70) | 0.90 | (0.84-0.95) | 0.93 | (0.89-0.98) |
| PCPS | 0.65 | (0.58-0.72) | 0.76 | (0.65-0.88) | 0.82 | (0.71-0.93) |
| DCCS | 0.56 | (0.48-0.64) | 0.69 | (0.55-0.84) | 0.86 | (0.76-0.95) |
| Data presented as means (95% confidence intervals). HC – Healthy Control; MCI – Mild Cognitive Impairment; AD – Alzheimer’s disease; ADAS-Cog13 - Alzheimer's Disease Assessment Scale-Cognitive Subscale 13; PCPS – NIH Toolbox Pattern Comparison Processing Speed Test; DCCS - NIH Toolbox Dimensional Change Card Sort |  |  |  |  |  |  |

### NIH Toolbox assessments of executive functioning decline as early as 6 months prior to cognitive decline measured by ADAS-Cog13

In light of the differences in variances between tests among patients, we next sought to determine whether longitudinal patterns between cognitive outcomes (declined, stable, improved) were correlated with cognitive testing trends. Among the AD patients within the GAINS cohort, we first explored differences between those who experienced a decline in cognition versus those who did not (both stable and improved patients). We did not observe any differences in demographic or clinical characteristics, including in the medications taken by the subjects during the study period (**Supplemental Table 3**). We did, however, notice a significant decline in executive functioning testing, in both NIH toolbox PCPS and DCCS, at the 3- and 6-month visits before a decline in cognition measured by the ADAS-Cog13 compared to all other timepoints where the ADAS-Cog13 testing was stable (**Figure 1a**). There were no significant differences in ADAS-Cog13 testing scores at these same time points. The differences from baseline score tests are visualized in **Figure 1b**.

**Figure 1:**
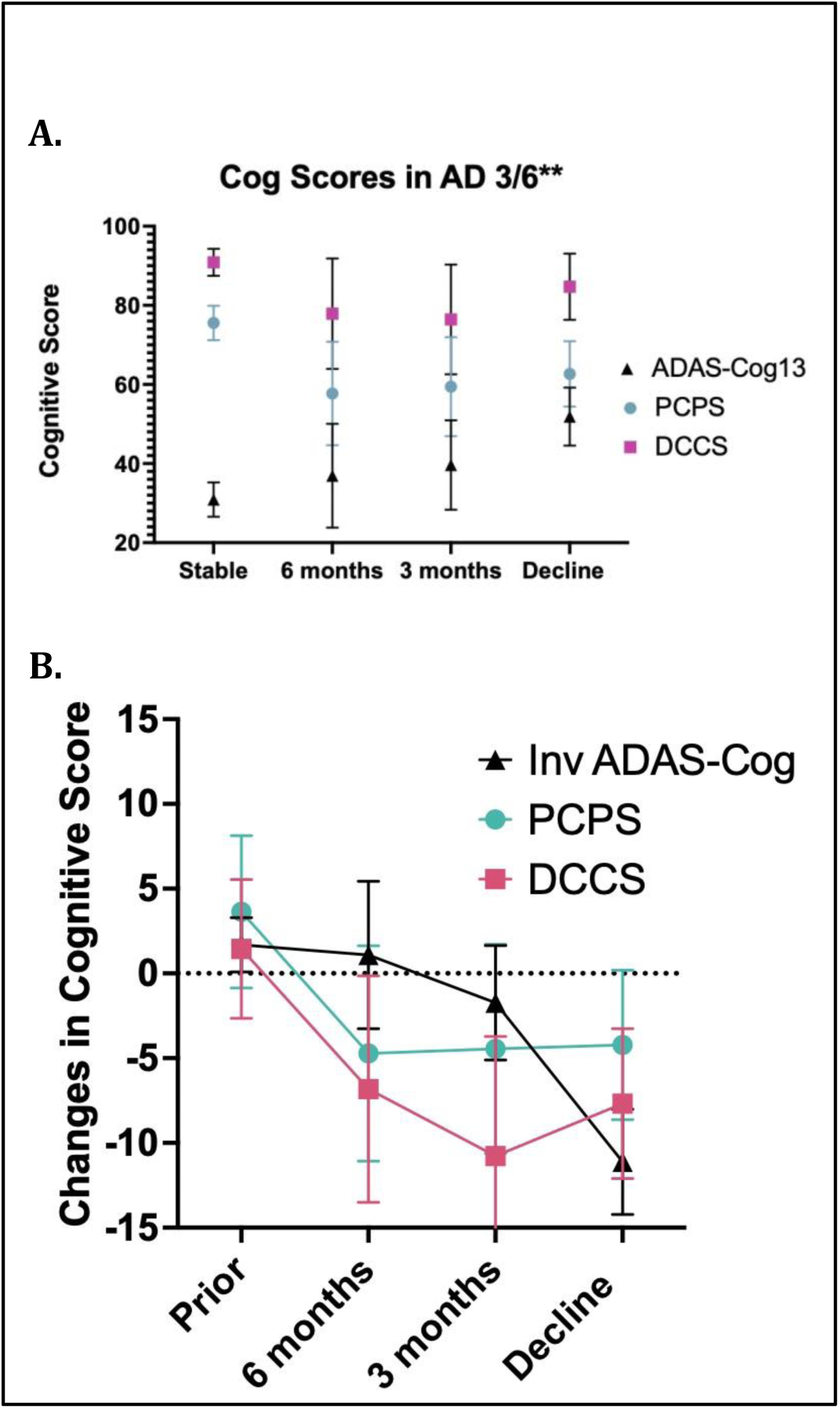
Clinical cognitive score testing among Alzheimer’s disease patients.

**Supplemental Table 3:**
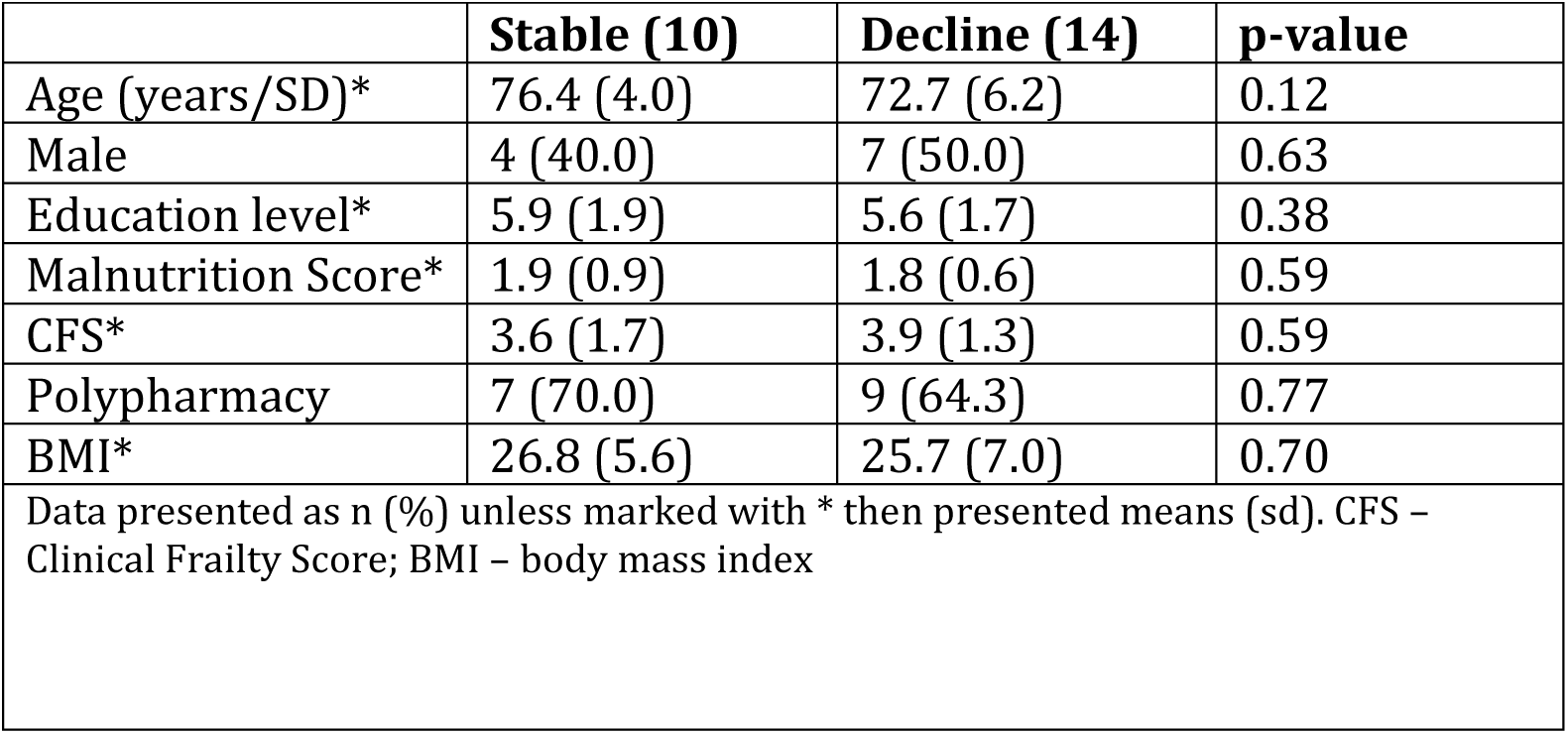
Clinical Characteristics between Alzheimer’s disease patients with and without cognitive decline.

We next performed univariate cox models predicting cognitive decline by ADAS-Cog for both NIH toolbox domains for PCPS and DCCS among AD patients. We used a 5-point decline in both NIH CB tests as predictor of general cognitive decline given the mean change at both 3 and 6 months prior in each test was about 5 points. Both in the crude and adjusted models, a 5-point change in PCPS or DCCS was associated with a 24% to 32% or 44% to 62% increased risk of cognitive decline respectively (**Table 4**). In the modeling, adjustments were made for age, sex, education, frailty, malnutrition and polypharmacy.

**Table 4:** Analysis of NIH Toolbox Cognitive Testing and Risk of Cognitive Decline.

|  |  |  |  |  |
| --- | --- | --- | --- | --- |
|  | **Crude** |  | **Adjusted †** |  |
|  | **Hazard Ratio**<br>**(95% CI)** | **P value** | **Hazard Ratio**<br>**(95% CI)** | **P value** |
| PCPS delta | **1.24 (1.08-1.43)** | **0.002** | **1.32 (1.08-1.60)** | **0.006** |
| DCCS delta | **1.44 (1.16-1.77)** | **0.001** | **1.62 (1.16-2.26)** | **0.005** |
| PCPS – NIH Toolbox Pattern Comparison Processing Speed Test; DCCS - NIH Toolbox Dimensional Change Card Sort; delta is for every 5-point decrease in score<br>† Adjusted for age, sex, education, frailty, malnutrition, and polypharmacy |  |  |  |  |

## DISCUSSION

In this study we found that as early as 6 months prior to a global cognitive decline, there were significant decreases in processing speed and executive functioning among AD patients using the NIH toolbox CB assessments for PCPS and DCCS. There was also a noted increase in the between-person variability from cHC to MIC to AD subjects using either the ADAS-Cog or NIH toolbox modules, however the within-group variability only increased between these groups in the ADAS-Cog assessments, with the greatest variability in among AD patients. A 5-point decrease in either PCPS and DCCS resulted in a greater than 30% increased risk of subsequent global cognitive decline. We propose here that these NIH toolbox assessments for executive functioning and processing speed may serve as a monitoring tool to predict which AD patients will go on to experience clinically significant cognitive decline.

Executive function comprises the higher-level cognitive skills used to control and coordinate other cognitive abilities and behaviors (12, 13). Deficits in executive functioning have been shown as one of the most useful cognitive markers for the early detection of AD (17), and declining performance can be detected 2-3 years before the diagnosis of AD (49). Executive dysfunction occurs in all stages of AD and has been linked to functional decline in activities of daily living (50, 51). Executive functioning is also one of the cognitive domains in AD that shows the greatest decline and can indicate a faster disease progression (52–54). The ADAS-Cog test does a poor job of testing for executive functioning. It was built based on the original ADAS-Cog (55), and the ADAS-Cog13 or ADAS-Cog-Modified had delayed word recall or number cancellation added in order to improve the tests responsiveness, to cover the range of mild to moderate AD (25). Others tried adding additional testing domains to the ADAS-Cog test to better cover executive functioning testing such as the ADAS-Cog-Exec (56) and ADAS-Cog-Plus (26). These additional testing modules are not universally used, with the ADAS-Cog still considered the gold standard (27) especially for assessing efficacy of AD treatments.

As we age, both executive functioning and processing speed decline, and this decline is linked to performance in learning and memory (57). The processing speed theory states that the age-related decline in processing speed is the fundamental mechanism with which memory declines with normal aging (58). However, executive functions and processing speed can differentially influence memory decline (57). Processing speed is thought to serve as the foundation for other cognitive processes (15) and it is associated with subsequent deficits in other cognitive domains such as working memory (59), attention (60) and memory (61). Disproportionate slowing of processing speed has been shown to be related to a faster decline in AD (18). Our finding is consistent with a predictive role of processing speed change: the NIH toolbox CB Comparison Processing Speed Test to assess processing speed in GAINS participants was sensitive to early cognitive change, with decreases in processing speed preceding global cognitive decline, with a 5 point drop in score increasing the risk of cognitive decline by greater than 30%, after adjustment.

Instead of expanding the ADAS-Cog to improve its sensitivity, it might be of greater benefit to predict cognitive decline by periodically test executive functioning with other standard measures. The NIH toolbox CB is a set of brief measures assessing cognitive domains including executive functioning studies (19). Subjects that are part of a clinical trial could self-administer these tests as an alternative to the ADAS-Cog, which takes upwards of 45 minutes to administer and needs to be done in-person by trained staff (27). Remote digital cognitive testing is now being shown to be an accurate method to detect cognitive impairment (62–64). A robust tool with a 3 to 6-month predictive window would offer patients and families a reasonable timeframe to make practical care arrangements such as changing living situation, finding caretakers, or applying for elder care benefits and services.

This study does have limitations. First, the majority of the GAINS cohort was without AD, which influences the within-group variance in the AD group. Testing NIH toolbox assessments for their ability to predict cognitive decline in a larger longitudinal cohort would strengthen the findings. Additionally, the GAINS cohort was mostly white and non-Hispanic, limiting the generalizability of our data. These limitations are balanced by the frequent in-person model to assess both ADAS-Cog and the NIH toolbox CB domains.

## Conclusion

Based on our findings, we would suggest that brief assessments of both executive functioning and processing speed may serve as a marker of subsequent global cognitive decline. This can be helpful clinically for planning the timing of treatments, interventions, or life management, especially if executive function and processing speed testing is potentially more rapid than a comprehensive cognitive testing session, and more feasibly performed frequently. Tools such as PCPS and DCCS could also help researchers as well as clinicians with monitoring efficacy of AD treatments as part of clinical trials of disease-modifying therapy.

## AUTHOR CONTRIBUTIONS

JPH (Data curation; Conceptualization; Formal analysis; Investigation; Project administration; Funding acquisition; Writing – original draft); AMB (Methodology; Writing – review & editing); SNO (Supervision; Data curation; Writing – review & editing); PD (Supervision; Data curation; Writing – review & editing); PMM (Data acquisition; Data curation; Writing – review & editing); IS (Data acquisition; Writing – review & editing); LR (Data acquisition; Writing – review & editing); YSL (Resources; Writing – review & editing); VB (Conceptualization; Formal analysis; Methodology; Writing – review & editing); BAM (Conceptualization; Writing – review & editing).

## ACKNOWLEDEMENTS

We would like to thank the administration and staff from the Clinical Research Center here at UMass Medical Center and the Center for Clinical and Translational Sciences at UMass Chan Medical School for clinical facilities that supported the GAINS cohort.

## DATA AVAILABILITY

The data supporting the findings of this study are available on request from the corresponding author. The data are not publicly available due to privacy or ethical restrictions.

## REFERENCES

1. Rajan KB, Weuve J, Barnes LL, McAninch EA, Wilson RS, Evans DA. 2021. Population estimate of people with clinical Alzheimer’s disease and mild cognitive impairment in the United States (2020-2060). Alzheimers Dement 17:1966–1975.

2. Schindler SE, Li Y, Buckles VD, Gordon BA, Benzinger TLS, Wang G, Coble D, Klunk WE, Fagan AM, Holtzman DM, Bateman RJ, Morris JC, Xiong C. 2021. Predicting Symptom Onset in Sporadic Alzheimer Disease With Amyloid PET. Neurology 97:e1823–e1834.

3. Koscik RL, Betthauser TJ, Jonaitis EM, Allison SL, Clark LR, Hermann BP, Cody KA, Engle JW, Barnhart TE, Stone CK, Chin NA, Carlsson CM, Asthana S, Christian BT, Johnson SC. 2020. Amyloid duration is associated with preclinical cognitive decline and tau PET. Alzheimers Dement (Amst) 12:e12007.

4. Insel PS, Donohue MC, Berron D, Hansson O, Mattsson-Carlgren N. 2021. Time between milestone events in the Alzheimer’s disease amyloid cascade. Neuroimage 227:117676.

5. Xie J, Brayne C, Matthews FE. 2008. Survival times in people with dementia: analysis from population based cohort study with 14 year follow-up. Bmj 336:258–62.

6. Adak S, Illouz K, Gorman W, Tandon R, Zimmerman EA, Guariglia R, Moore MM, Kaye JA. 2004. Predicting the rate of cognitive decline in aging and early Alzheimer disease. Neurology 63:108–14.

7. Cosentino S, Scarmeas N, Helzner E, Glymour MM, Brandt J, Albert M, Blacker D, Stern Y. 2008. APOE epsilon 4 allele predicts faster cognitive decline in mild Alzheimer disease. Neurology 70:1842–9.

8. Steenland K, Zhao L, Goldstein F, Cellar J, Lah J. 2014. Biomarkers for predicting cognitive decline in those with normal cognition. J Alzheimers Dis 40:587–94.

9. Gunes S, Aizawa Y, Sugashi T, Sugimoto M, Rodrigues PP. 2022. Biomarkers for Alzheimer’s Disease in the Current State: A Narrative Review. Int J Mol Sci 23.

10. Wang H, Sun M, Li W, Liu X, Zhu M, Qin H. 2023. Biomarkers associated with the pathogenesis of Alzheimer’s disease. Front Cell Neurosci 17:1279046.

11. Henneges C, Reed C, Chen YF, Dell’Agnello G, Lebrec J. 2016. Describing the Sequence of Cognitive Decline in Alzheimer’s Disease Patients: Results from an Observational Study. J Alzheimers Dis 52:1065–80.

12. Memory. Aging Centerand Aging Center, UCSF Weill Institute for Neurosciences. Executive functions. Available at: https://memory.ucsf.edu/symptoms/executive-functions. Accessed June 6, 2024.

13. Guarino A, Favieri F, Boncompagni I, Agostini F, Cantone M, Casagrande M. 2018. Executive Functions in Alzheimer Disease: A Systematic Review. Front Aging Neurosci 10:437.

14. Ebaid D, Crewther SG, MacCalman K, Brown A, Crewther DP. 2017. Cognitive Processing Speed across the Lifespan: Beyond the Influence of Motor Speed. Front Aging Neurosci 9:62.

15. Sliwinski M, Buschke H. 1999. Cross-sectional and longitudinal relationships among age, cognition, and processing speed. Psychol Aging 14:18–33.

16. Rabinovici GD, Stephens ML, Possin KL. 2015. Executive dysfunction. Continuum (Minneap Minn) 21:646–59.

17. Crowell TA, Luis CA, Vanderploeg RD, Schinka JA, Mullan M. 2002. Memory Patterns and Executive Functioning in Mild Cognitive Impairment and Alzheimer’s Disease. Aging, Neuropsychology, and Cognition 9:288–297.

18. Parikh M, Hynan LS, Weiner MF, Lacritz L, Ringe W, Cullum CM. 2014. Single neuropsychological test scores associated with rate of cognitive decline in early Alzheimer disease. Clin Neuropsychol 28:926–40.

19. Weintraub S, Dikmen SS, Heaton RK, Tulsky DS, Zelazo PD, Slotkin J, Carlozzi NE, Bauer PJ, Wallner-Allen K, Fox N, Havlik R, Beaumont JL, Mungas D, Manly JJ, Moy C, Conway K, Edwards E, Nowinski CJ, Gershon R. 2014. The cognition battery of the NIH toolbox for assessment of neurological and behavioral function: validation in an adult sample. J Int Neuropsychol Soc 20:567–78.

20. Zelazo PD, Anderson JE, Richler J, Wallner-Allen K, Beaumont JL, Conway KP, Gershon R, Weintraub S. 2014. NIH Toolbox Cognition Battery (CB): validation of executive function measures in adults. J Int Neuropsychol Soc 20:620–9.

21. Carlozzi NE, Beaumont JL, Tulsky DS, Gershon RC. 2015. The NIH Toolbox Pattern Comparison Processing Speed Test: Normative Data. Archives of Clinical Neuropsychology 30:359–368.

22. Warren SL, Reid E, Whitfield P, Helal AM, Abo Hamza EG, Tindle R, Moustafa AA, Hamid MS. 2024. Cognitive and behavioral abnormalities in individuals with Alzheimer’s disease, mild cognitive impairment, and subjective memory complaints. Current Psychology 43:800–810.

23. Schrag A, Schott JM. 2012. What is the clinically relevant change on the ADAS-Cog? J Neurol Neurosurg Psychiatry 83:171–3.

24. Karcher H, Savelieva M, Qi L, Hummel N, Caputo A, Risson V, Capkun G, Alzheimer’s Disease Neuroimaging I. 2020. Modelling Decline in Cognition to Decline in Function in Alzheimer’s Disease. Curr Alzheimer Res 17:635–657.

25. Mohs RC, Knopman D, Petersen RC, Ferris SH, Ernesto C, Grundman M, Sano M, Bieliauskas L, Geldmacher D, Clark C, Thal LJ. 1997. Development of cognitive instruments for use in clinical trials of antidementia drugs: additions to the Alzheimer’s Disease Assessment Scale that broaden its scope. The Alzheimer’s Disease Cooperative Study. Alzheimer Dis Assoc Disord 11 Suppl 2:S13–21.

26. Skinner J, Carvalho JO, Potter GG, Thames A, Zelinski E, Crane PK, Gibbons LE. 2012. The Alzheimer’s Disease Assessment Scale-Cognitive-Plus (ADAS-Cog-Plus): an expansion of the ADAS-Cog to improve responsiveness in MCI. Brain Imaging Behav 6:489–501.

27. Kueper JK, Speechley M, Montero-Odasso M. 2018. The Alzheimer’s Disease Assessment Scale-Cognitive Subscale (ADAS-Cog): Modifications and Responsiveness in Pre-Dementia Populations. A Narrative Review. J Alzheimers Dis 63:423–444.

28. Rubenstein LZ, Harker JO, Salvà A, Guigoz Y, Bruno Vellas B. 2001. Screening for Undernutrition in Geriatric Practice: Developing the Short-Form Mini-Nutritional Assessment (MNA-SF). J Gerontol A Biol Sci Med Sci 56A:M366-72.

29. Saarela RK, Lindroos E, Soini H, Hiltunen K, Muurinen S, Suominen MH, Pitkala KH. 2016. Dentition, nutritional status and adequacy of dietary intake among older residents in assisted living facilities. Gerodontology doi:10.1111/ger.12144.

30. Guigoz Y. 2006. The Mini Nutritional Assessment (MNA) review of the literature--What does it tell us? J Nutr Health Aging 10:485–7.

31. Rockwood K, Song X, MacKnight C, Bergman H, Hogan DB, McDowell I, Mitnitski A. 2005. A global clinical measure of fitness and frailty in elderly people. CMAJ 173:489–95.

32. Connor DJ, Sabbagh MN. 2008. Administration and scoring variance on the ADAS-Cog. J Alzheimers Dis 15:461–4.

33. Gershon RC, Wagster MV, Hendrie HC, Fox NA, Cook KF, Nowinski CJ. 2013. NIH Toolbox for Assessment of Neurological and Behavioral Function. Neurology 80:S2–6.

34. Anonymous. NIH Toolbox Cognitive Battery (NIHTB-CB): The NIHTB Pattern Comparison Processing Speed Test. doi:10.1017/S1355617714000319.

35. Matthews HP, Korbey J, Wilkinson DG, Rowden J. 2000. Donepezil in Alzheimer’s disease: eighteen month results from Southampton Memory Clinic. Int J Geriatr Psychiatry 15:713–20.

36. Aisen PS, Schafer KA, Grundman M, Pfeiffer E, Sano M, Davis KL, Farlow MR, Jin S, Thomas RG, Thal LJ. 2003. Effects of rofecoxib or naproxen vs placebo on Alzheimer disease progression: a randomized controlled trial. Jama 289:2819–26.

37. Le Bars PL, Kieser M, Itil KZ. 2000. A 26-week analysis of a double-blind, placebo-controlled trial of the ginkgo biloba extract EGb 761 in dementia. Dement Geriatr Cogn Disord 11:230–7.

38. Masnoon N, Shakib S, Kalisch-Ellett L, Caughey GE. 2017. What is polypharmacy? A systematic review of definitions. BMC Geriatr 17:230.

39. Jackson MA, Jeffery IB, Beaumont M, Bell JT, Clark AG, Ley RE, O’Toole PW, Spector TD, Steves CJ. 2016. Signatures of early frailty in the gut microbiota. Genome Med 8:8.

40. Milani C, Ticinesi A, Gerritsen J, Nouvenne A, Lugli GA, Mancabelli L, Turroni F, Duranti S, Mangifesta M, Viappiani A, Ferrario C, Maggio M, Lauretani F, De Vos W, van Sinderen D, Meschi T, Ventura M. 2016. Gut microbiota composition and Clostridium difficile infection in hospitalized elderly individuals: a metagenomic study. Sci Rep 6:25945.

41. Haran JP, Bucci V, Dutta P, Ward D, McCormick B. 2018. The nursing home elder microbiome stability and associations with age, frailty, nutrition, and physical location. J Med Microbiol 67:40–51.

42. Harris PA, Taylor R, Thielke R, Payne J, Gonzalez N, Conde JG. 2009, Apr. Research electronic data capture (REDCap) - A metadata-driven methodology and workflow process for providing translational research informatics support. J Biomed 42:377–81.

43. Guerin O, Soto ME, Brocker P, Robert PH, Benoit M, Vellas B, Group. RF. 2005. Nutritional status assessment during Alzheimer’s disease: results after one year (the REAL French Study Group). J Nutr Health Aging 9:81–4.

44. Buchman AS, Schneider JA, Leurgans S, Bennett DA. 2008. Physical frailty in older persons is associated with Alzheimer disease pathology. Neurology 71:499–504.

45. Buchman AS, Boyle PA, Wilson RS, Tang Y, Bennett DA. 2007. Frailty is associated with incident Alzheimer’s disease and cognitive decline in the elderly. Psychosom Med 69:483–9.

46. Meijers JM, Schols JM, Halfens RJ. 2014. Malnutrition in care home residents with dementia. J Nutr Health Aging 18:595–600.

47. Boulos C, Salameh P, Barberger-Gateau P. 2016. Malnutrition and frailty in community dwelling older adults living in a rural setting. Clin Nutr 35:138–43.

48. Salthouse TA. 2007. Implications of within-person variability in cognitive and neuropsychological functioning for the interpretation of change. Neuropsychology 21:401–11.

49. Grober E, Hall CB, Lipton RB, Zonderman AB, Resnick SM, Kawas C. 2008. Memory impairment, executive dysfunction, and intellectual decline in preclinical Alzheimer’s disease. J Int Neuropsychol Soc 14:266–78.

50. Tekin S, Fairbanks LA, O’Connor S, Rosenberg S, Cummings JL. 2001. Activities of daily living in Alzheimer’s disease: neuropsychiatric, cognitive, and medical illness influences. Am J Geriatr Psychiatry 9:81–6.

51. Skurla E, Rogers JC, Sunderland T. 1988. Direct assessment of activities of daily living in Alzheimer’s disease. A controlled study. J Am Geriatr Soc 36:97–103.

52. Chen P, Ratcliff G, Belle SH, Cauley JA, DeKosky ST, Ganguli M. 2001. Patterns of cognitive decline in presymptomatic Alzheimer disease: a prospective community study. Arch Gen Psychiatry 58:853–8.

53. Tosto G, Gasparini M, Brickman AM, Letteri F, Renie R, Piscopo P, Talarico G, Canevelli M, Confaloni A, Bruno G. 2015. Neuropsychological predictors of rapidly progressive Alzheimer’s disease. Acta Neurol Scand 132:417–22.

54. Zhao Q, Zhou B, Ding D, Teramukai S, Guo Q, Fukushima M, Hong Z. 2014. Cognitive decline in patients with Alzheimer’s disease and its related factors in a memory clinic setting, Shanghai, China. PLoS One 9:e95755.

55. Rosen WG, Mohs RC, Davis KL. 1984. A new rating scale for Alzheimer’s disease. Am J Psychiatry 141:1356–64.

56. Jacobs DM, Thomas RG, Salmon DP, Jin S, Feldman HH, Cotman CW, Baker LD. 2020. Development of a novel cognitive composite outcome to assess therapeutic effects of exercise in the EXERT trial for adults with MCI: The ADAS-Cog-Exec. Alzheimers Dement (N Y) 6:e12059.

57. Saikia B, Tripathi R. 2024. Executive Functions, Processing Speed, and Memory Performance: Untangling the Age-related Effects. Journal of Psychiatry Spectrum 3:12–19.

58. Salthouse TA. 1996. The processing-speed theory of adult age differences in cognition. Psychol Rev 103:403–28.

59. Chiaravalloti ND, Christodoulou C, Demaree HA, DeLuca J. 2003. Differentiating simple versus complex processing speed: influence on new learning and memory performance. J Clin Exp Neuropsychol 25:489–501.

60. Mayes SD, Calhoun SL. 2007. Learning, attention, writing, and processing speed in typical children and children with ADHD, autism, anxiety, depression, and oppositional-defiant disorder. Child Neuropsychol 13:469–93.

61. Baudouin A, Clarys D, Vanneste S, Isingrini M. 2009. Executive functioning and processing speed in age-related differences in memory: contribution of a coding task. Brain Cogn 71:240–5.

62. Berron D, Olsson E, Andersson F, Janelidze S, Tideman P, Düzel E, Palmqvist S, Stomrud E, Hansson O. 2024. Remote and unsupervised digital memory assessments can reliably detect cognitive impairment in Alzheimer’s disease. Alzheimers Dement 20:4775–4791.

63. Boots EA, Frank RD, Fan WZ, Christianson TJ, Kremers WK, Stricker JL, Machulda MM, Fields JA, Hassenstab J, Graff-Radford J, Vemuri P, Jack CR, Knopman DS, Petersen RC, Stricker NH. 2024. Continuous Associations between Remote Self-Administered Cognitive Measures and Imaging Biomarkers of Alzheimer’s Disease. The Journal of Prevention of Alzheimer’s Disease doi:10.14283/jpad.2024.99.

64. Butler J, Watermeyer TJ, Matterson E, Harper EG, Parra-Rodriguez M. 2024. The development and validation of a digital biomarker for remote assessment of Alzheimer’s diseases risk. DIGITAL HEALTH 10:20552076241228416.

